# A survey of Paediatric Radiology Artificial Intelligence

**DOI:** 10.1101/2024.09.19.24313885

**Authors:** Brendan S Kelly, Simon M Clifford, Conor S Judge, Stephanie M Bollard, Gerard M Healy, Hannah Hughes, Gabrielle C Colleran, J.E. Rod, Edward H Lee, Laura M Prolo, Kristen W Yeom, Aonghus Lawlor, Ronan P Killeen

## Abstract

**Background:** Artificial intelligence (AI) applications in paediatric radiology present unique challenges due to diverse anatomy and physiology across age groups. Advancements in AI algorithms, particularly deep learning techniques, show promise in improving diagnostic accuracy.

**Objectives:** To survey trends in AI research in paediatric radiology. To evaluate use cases, tasks, research methodologies and underlying data. To identify potential biases and future directions.

**Methods:** A systematic search of paediatric radiology AI studies published from 2015 to 2021 was conducted following the PRISMA guidelines and the Cochrane Collaboration Handbook. The search included papers utilizing AI techniques for radiological diagnosis or intervention in patients aged under 18. Narrative synthesis was used due to methodological heterogeneity.

**Results:** A total of 292 articles were included, with an increasing annual trend in the number of published articles. Neuroradiology and musculoskeletal radiology were the most common subspecialties. MRI was the dominant imaging modality, with segmentation and classification as the most common tasks. Retrospective cohort studies constituted the majority of research designs. Data quality and quantity varied, as did the choice of research design, data sources, and evaluation metrics.

**Conclusions:** AI literature in paediatric radiology shows rapid growth, with advancements in various subspecialties and tasks. However, potential biases and data quality issues highlight the need for rigorous research design and evaluation to ensure the generalisability and reliability of AI models in clinical practice. Future research should focus on addressing these biases and improving the robustness of AI applications in paediatric radiology.

## Introduction

AI applications in radiology are anticipated to significantly impact the entire specialty [1]. As an umbrella term, AI refers to a computer’s ability to mimic human behaviour, while Machine Learning (ML), a subset of AI, is generally understood as the capacity to learn without being explicitly programmed. AI algorithms, especially those employing deep learning techniques, efficiently processed and learned from large datasets, accounting for the extensive variability and accurately identifying subtle differences in data. As the volume of data collected for medical imaging increased, so too did the opportunities for large-scale analysis with both classical machine learning techniques and radiomics, which involved the extraction of quantitative features [2], and more recently, computer vision methods leveraging deep learning techniques [3]. Recent papers suggest that human expert-level performance is now attainable by AI models[4], which heightened interest in the topic [5], leading to a surge in the literature [6, 7].

Paediatric radiology presents unique challenges, making AI particularly interesting in this field[8]. There is a global shortage of paediatric subspecialists in radiology, and 97% of radiology departments overall unable to meet imaging demands within contracted hours [9]. Additionally, AI has the potential to minimise radiation exposure by optimising image acquisition and reducing the need for repeat scans, crucial for this vulnerable demographic [10]. This is especially important considering that in general radiology demand for cross-sectional imaging is increasing 10% annually [9]. There is much diversity in anatomy and physiology across various age groups, from neonates to adolescents, which necessitates precise and adaptable image interpretation. Other factors that motivate the use of AI in paediatric imaging include the wide variation in normal and the plethora of rare syndromic and genetic diseases[10]. Children are also more radiosensitive than adults and as such ionising radiation must be justified to a higher standard with fewer radiologic studies available to analyse as a direct result[10]. Similarly MRI may need the use of general anesthetic or sedation, again raising the standard of justification[11]. Movement artifacts may be more of an issue across modalities and the use of low dose techniques may reduce the signal-to-noise ratio which could impact machine learning.

Certain paediatric-specific use cases were particularly suitable for AI[8], with tasks such as bone age estimation[12] as likely candidates for automation. However, a delay occurred between the initial excitement surrounding radiology AI and the implementation of tools into clinical practice. This gap between literature and practice was described as the “AI chasm” [13]. Specific challenges, such as patient diversity, a broad range of normal variation across ages, and the higher threshold for imaging in paediatrics, made the situation more complex than initially anticipated. Despite these challenges, the clinical application of AI in paediatric radiology remains an active area of research, with new advancements continuously emerging[14]. Multidisciplinary collaboration among researchers, clinicians, and industry partners will be necessary to overcome these challenges and realize the full potential of AI in paediatric radiology[14]. A key aspect necessary for an effective multidisciplinary collaboration is to identify trends in the relevant research methods that could promote a common understanding and resolve potential inconsistencies.

Therefore, the aims of the present review is to provide a survey that identifies relevant methodological trends of published studies that discuss the use of AI in paediatric radiology by performing a systematic search. The methodological trends of interest includes (i) an evaluation of the clinical use cases, study methods and AI tasks, (ii) a comprehensive review of data quantity-quality and, (iii) evaluate the presence of potential biases introduced by the chosen methods and evaluate their potential impact on study outcomes.

A survey review addressing the above aims could act as a reference point for researchers in the field [15]. In addition, it could facilitate multidisciplinary collaboration among a variety of stakeholders by offering valuable insights into the state of AI applications in paediatric radiology. Finally, a survey review could offer a perspective about how current AI research is helping to address paediatric radiology challenges and provide insights that help guide future research.

## Methods

The authors adhered to the Preferred Reporting Items for Systematic Review and Meta-Analysis guidelines (PRISMA)[16] and the Cochrane Collaboration Handbook [17], conducting a systematic review of all paediatric radiology AI studies published from 2015 to 2021 in accordance with a published protocol[18] of a prospectively registered review (PROSPERO: CRD42020154790). Full details can be found in the protocol. The need for ethical approval was waived at the departmental level.

### Inclusion Criteria

Hospital-based human studies that employed AI techniques to aid in the care of patients aged under 18 through radiological diagnosis or intervention were included. Studies that utilised machine learning techniques[19] to complete segmentation, identification, classification, or prediction tasks based on radiographic, Computed Tomography (CT), Magnetic Resonance (MR), Ultrasound (US), or nuclear medicine/molecular or hybrid imaging techniques were also included.

### Exclusion Criteria

Functional MRI (fMRI) papers were not included, as the techniques used in the computer analysis of fMRI data are quite distinct from the computer vision-based tasks that were the subject of this review. Papers solely for use in radiation therapy were also excluded. Non-human or phantom studies were not considered. Papers that included participants both over and under the age of 18 but did not separate results based on age to allow for separate analysis were excluded.

### Electronic search

Electronic searches were performed on MEDLINE (Pubmed) and EMBASE from 1 January 2015 until 31 December 2021. Zotero served as the reference manager, and the Revtools package on R was used to eliminate duplicate records[20]. The search was conducted in English. The search terms used can be found in Online Supplementary Material 1. The Artificial Intelligence and Radiology terms were combined using the AND operator, with the addition of the paediatric terms also combined using the AND operator.

### Selection and analysis of papers

The titles and abstracts of studies were reviewed to identify clinical radiological artificial intelligence studies for inclusion or exclusion. Studies with insufficient information to determine the use of AI computer vision methods were also included for full-text review. A full-text review was then performed, to confirm eligibility for inclusion in the final systematic review. Abstract, title, and full-text review were performed by B.K. and S.B. Disagreements were resolved by consensus or, if necessary, by a third reviewer (R.K.). Data extraction was undertaken by three radiologists, all of whom were nationally certified and had a research interest in artificial intelligence (B.K., S.C., and G.H.). Additionally, B.K. is a PhD candidate in radiology artificial intelligence, G.H. holds an MD in artificial intelligence, and S.C. is a specialist paediatric radiologist. Additional data extraction for the revision was carried out by H.H. and J.R. whose work was reviewed by B.K. and S.C. H.H. is a radiology resident with prior publications in bibliometric analysis of AI in radiology, while J.R. is a physician and epidemiologist with experience in AI, data science and knowledge synthesis projects. This process is summarised in a PRISMA flowchart (Figure 1).

**Figure 1.**
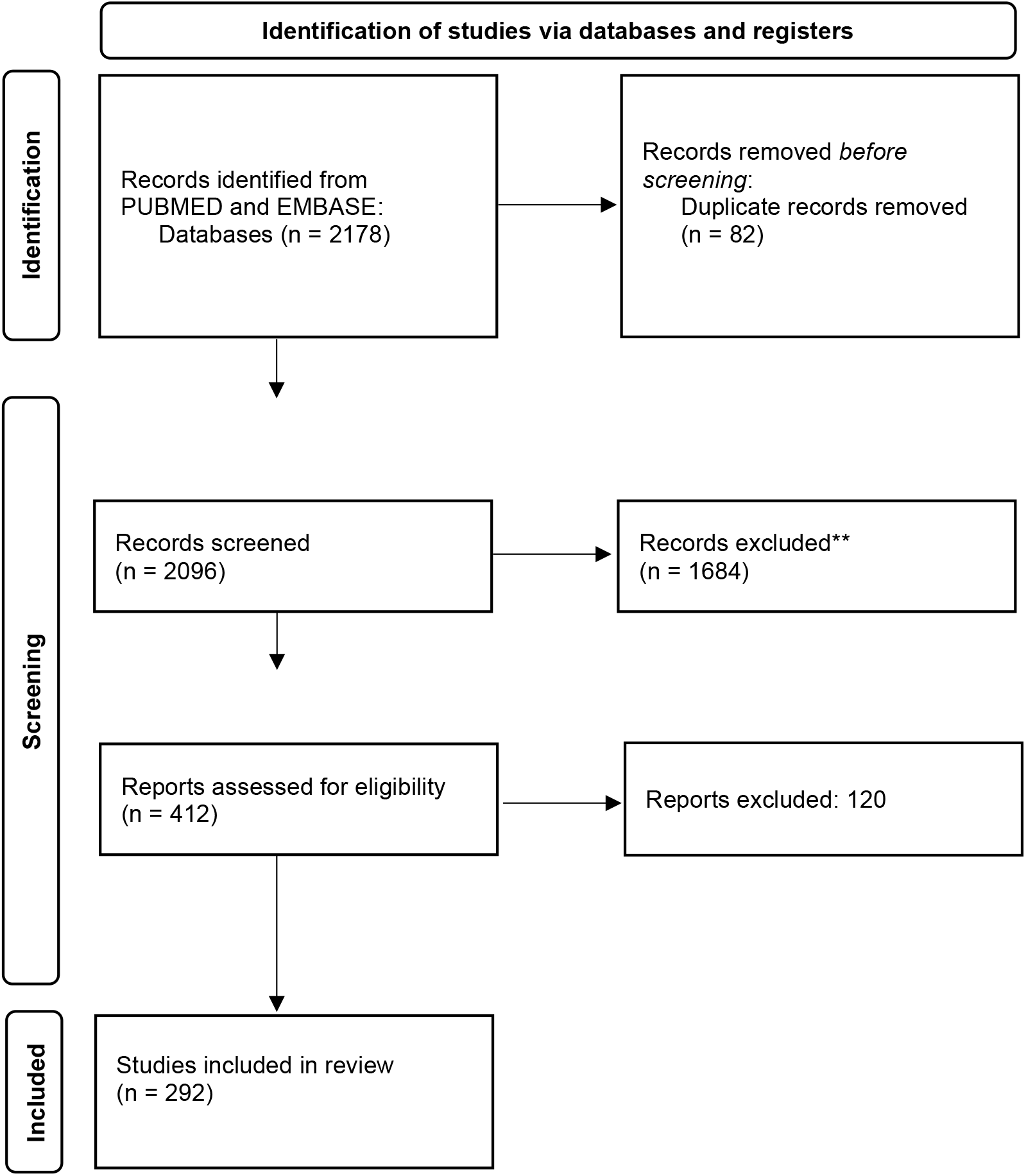
Prisma Flow Diagram. The PRISMA Flow diagram of papers included in our review

### Data Analysis

Data were analysed primarily using pivot tables and built-in exploratory analysis tools in Microsoft Excel. Due to the heterogeneity of employed methodologies, the synthesis and analysis of the extracted data was performed in a narrative format. This step was performed for each of the aims of the review. Taking into account the predicted heterogeneity of the methodologies [18], a limited bias assessment was conducted focussing on the Data, Algorithm (task) Training and Output (DATO) methods[21].

## Results

### Included papers

Our search yielded 2178 results. Titles and abstracts were screened by two reviewers and 412 full texts were reviewed. 292 articles were included for analysis. Details can be found in Figure 1. Online Supplementary Material 1 lists all included papers. There was an increasing trend in the number of included articles year on year with 21, 20, 22, 32, 48, 71, 78 articles respectively from 2015 to 2021 (Figures 2 and 3). The most common subspecialty each year was neuroradiology followed by musculoskeletal radiology (Figure 2). The diversity of subspecialtites increased year on year (Figure 2). A similar trend was observed in the choice of modality used, which was dominated by MRI (138, 47%) followed by radiography (86, 29%) (Figure 3). 34% of included papers had an author from the USA. Apart from the USA, researchers from China (21%), the UK (8%), Brazil (8%) and Canada (%) made up the majority of authorship.

**Figure 2.**
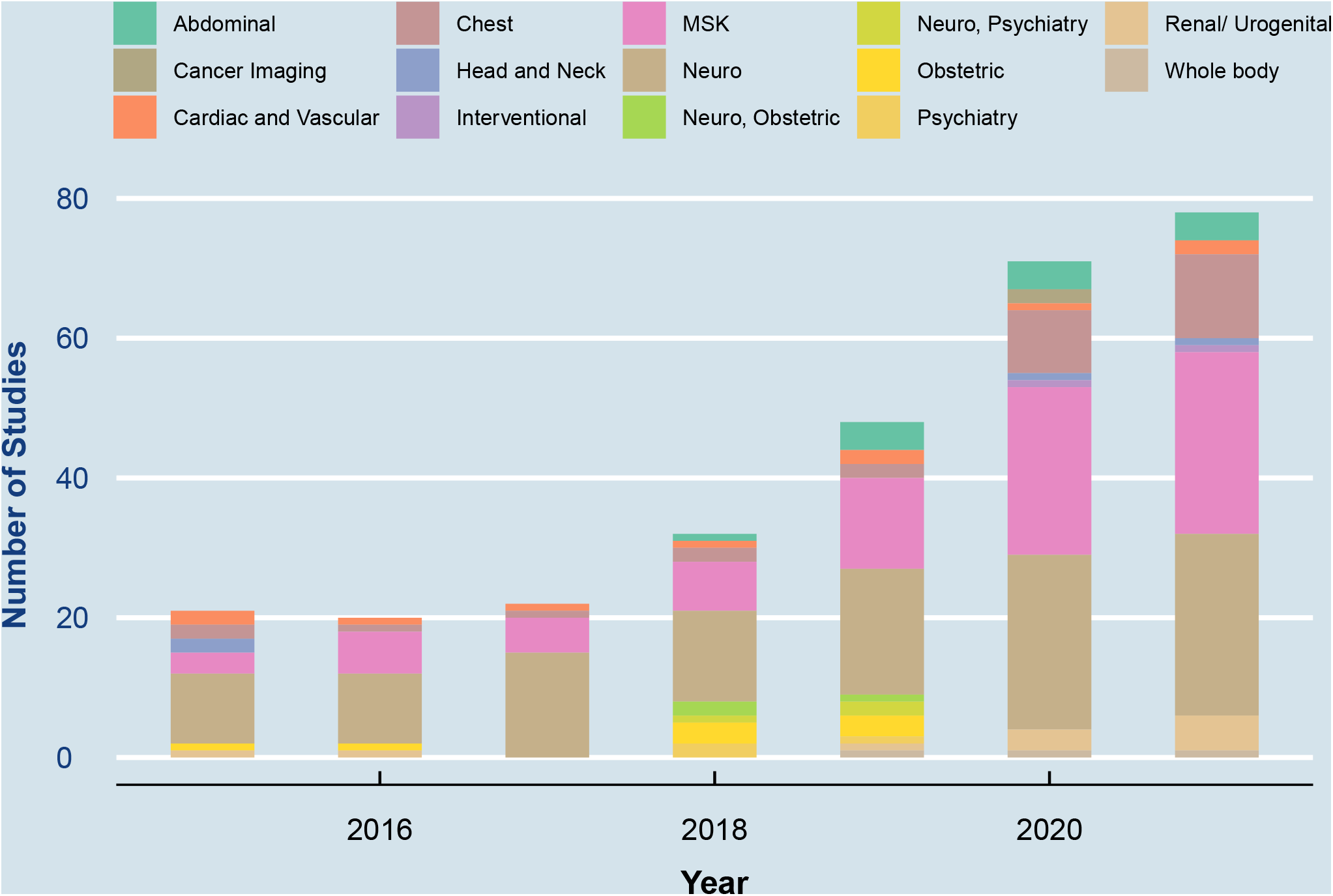
Paediatric Artificial Intelligence Articles by clinical area and year. Histogram by year of articles included in our review, colour denotes subspecialty

**Figure 3.**
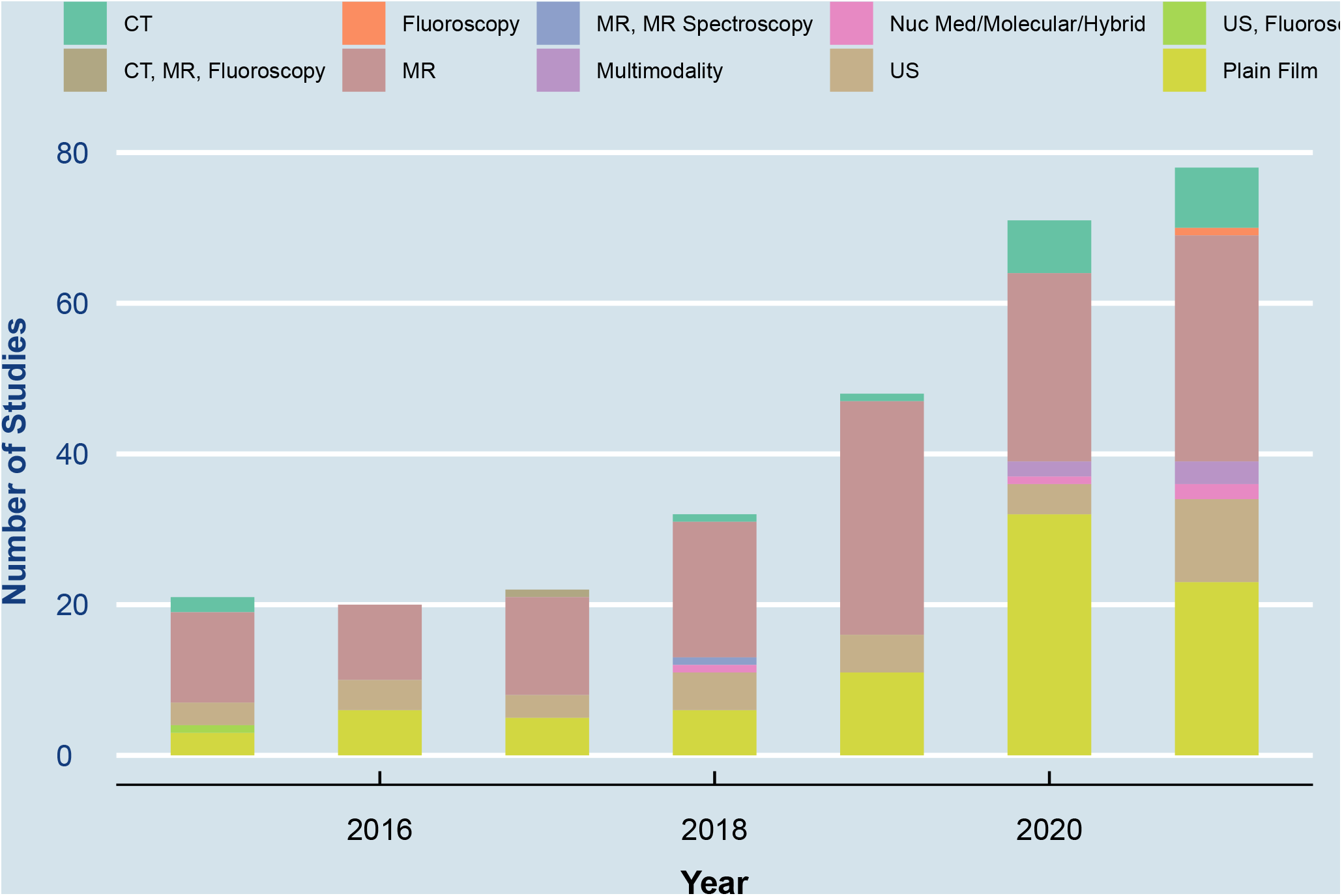
Paediatric Artificial Intelligence Articles by modality and year. Modality investigated by year. Colour denotes modality.

### Survey of Trends

#### Use cases, tasks and methods

The most common use case, seen in 52 (18%) of studies was neurodevelopmental disorders. This was followed by bone age 45 (15%) and 39 (13%) segmentation of normal anatomy, followed by classification and identification tasks. The most common clinical questions asked by subspecialty are summarised in Table 1. The most common task undertaken was segmentation 116 (40%), classification 79 (27%), followed by identification/localisation 37 (13%) Table 2. Support vector machines were the most commonly used algorithm for classification (24, 30%) followed by custom Convolutional Neural Networks (9, 11%). For segmentation tasks by far the most popular model was U Net 32 (28%).

**Table 1.** Most common clinical questions. Legend Table presents the most common search terms by subspecialty, note the total is less than 292 as infrequent entries are omitted for readability.

| Subspeciality | Abdomi<br>nal | Cardiac<br>and<br>Vascula<br>r | Chest | Interve<br>ntional | MSK | Neuro | Obstetri<br>c | Psychiat<br>ry | Renal/<br>Urogeni<br>tal | Grand<br>Total |
| --- | --- | --- | --- | --- | --- | --- | --- | --- | --- | --- |
| Clinical Use Case |  |  |  |  |  |  |  |  |  |  |
| Asthma, Small Airway disease |  |  | 4 |  |  |  |  |  |  | 8 |
| Bone Age |  |  |  |  | 40 | 4 |  |  |  | 48 |
| Brain age |  |  |  |  |  | 11 |  |  |  | 15 |
| Brain development |  |  |  |  |  | 40 | 3 | 3 |  | 50 |
| Cancer Imaging | 2 |  |  |  |  | 20 |  |  |  | 26 |
| Cardiomyopathy |  | 2 |  |  |  |  |  |  |  | 6 |
| Congenital heart disease |  | 5 | 1 |  |  |  |  |  |  | 10 |
| DDH |  |  |  |  | 12 |  |  |  |  | 16 |
| Device ID |  |  | 1 | 1 |  |  |  |  |  | 6 |
| Inflammatory bowel disease | 2 |  |  |  |  |  |  |  |  | 6 |
| Intussusception | 2 |  |  |  |  |  |  |  |  | 6 |
| Neonatal respiratory morbidity |  |  | 3 |  |  |  |  |  |  | 7 |
| Pneumonia |  |  | 16 |  |  |  |  |  |  | 20 |
| Developmental Renal/ bladder abnormalities |  |  |  |  |  |  |  |  | 7 | 11 |
| Scoliosis |  |  |  |  | 13 |  |  |  |  | 17 |
| Segmentation | 3 | 1 | 2 | 1 | 5 | 19 | 4 |  | 1 | 40 |
| Toal for these common use cases | 9 | 8 | 27 | 2 | 70 | 94 | 7 | 3 | 8 | 232 |
| Overall total | 13 | 10 | 29 | 2 | 84 | 117 | 8 | 3 | 11 | 281 |

**Table 2.**
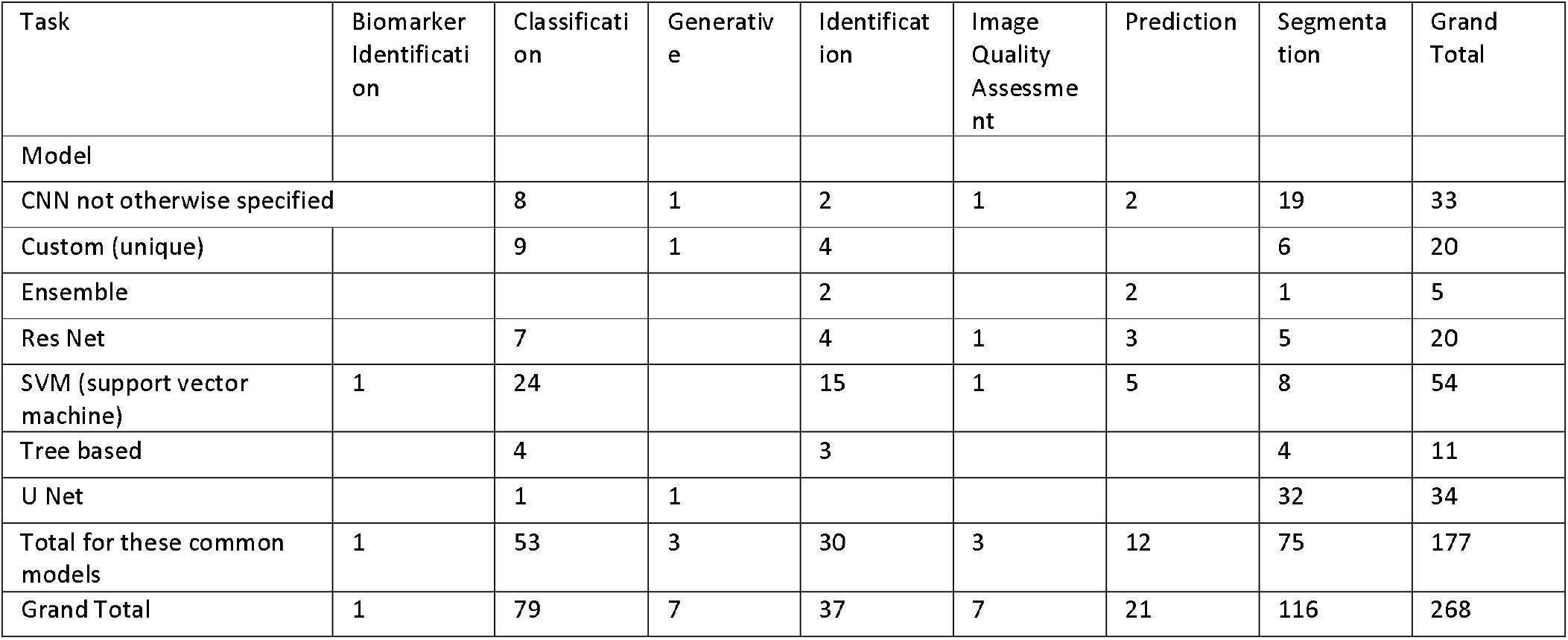
Most common AI methods. Legend Table presents the three most common algorithms used for each task, note the total is less than 292 as infrequent entries are omitted for readability.

86% (252) of studies were retrospective cohort studies. Ground truth was determined exclusively by the radiologic report in 90 31% of cases, and by an alternative specialist report on radiologic images in 34 12%. By an alternative imaging modality in 69 (24%) and by chronological age in 25 (9%). Inclusion criteria were explained in 265 (91%) of papers. No power analysis or sample size calculation was reported in 272 (93%) of studies. In the majority (186, 56%) of cases significant manual preprocessing of the images to be analysed was required prior to the use of the algorithm. The primary performance comparison was a radiologist in 94 (32%) of cases and an alternative medical specialist in 35 (12%). A “state of the art model” was used as the performance comparison in 26 (9%) of cases and there was no performance comparison in a further 50 (17%). The remainder were a combination of pathology, other modalities and reconstruction techniques.

Accuracy was the most common evaluation metric for classification tasks (22/79 28%). DICE was used as the metric in 52/116 45% of segmentation studies.

##### Data

Data were collected from only one hospital in 193 studies (66%), public databases in 39 (13%), more than one hospital in 32 (11%). The median number of patients included was 115 (range 10 - 14,236). The median number of images included was 135 (range 10 - 14236). The median number of images per patient was 1.2.

Data Augmentation was undertaken in 114 (39%) of papers. Of these, simple augmentation methods (symmetric operations, zooming, adding noise etc.) accounted for 66%, while synthetic image generation (using generative adversarial networks etc.) was utilized in 10%.

The source code or model was made freely available in 57 cases (20%) and available “upon reasonable request” in 7 (3%). The data used is private or unavailable in 172 (59%), publically available in 54 (18%) and available “upon reasonable request” in 22 (8%). No method of explainability was offered in 75 (26%) papers.The most common form of explainability was heat maps or other saliency visualisation offered in 50 papers (26%).

#### Trends

Data were collected from more than one hospital in 20/21 (95%) of cases in 2015 but by 2021 this had increased to 45/78 58%. Data or code were made available open access in only 5% of papers in 2015 but 20% of papers in 2021. The proportion of papers offering no method of explainability decreased year on year from 18/21 86% to 12/78 (15%) in 2021.

## Discussion

### Overview

This survey paper aimed to investigate the use of artificial intelligence (AI) in paediatric radiology by examining the tasks undertaken, methods employed, potential biases, data quality, and trends in the field. The results demonstrated that AI has been successfully applied to various tasks in paediatric radiology, with neurodevelopmental disorders and bone age estimation being the most common use cases. The majority of studies were retrospective cohort studies, and segmentation was the most frequently performed task. The findings of this survey are consistent with the existing literature [7, 15, 22, 23], highlighting the growing interest in AI applications within paediatric radiology. The research however is concentrated on a small cluster of use cases and uses a limited palette of techniques. This allows for identification of future areas for research as well as areas that could be approaching saturation.

#### Use cases methods and tasks

The most common tasks were segmentation, classification, and identification/localization. Support vector machines and custom Convolutional Neural Networks were popular choices for classification tasks, while U-Net dominated segmentation tasks. Accuracy and DICE were the most common evaluation metrics used. There is a strong focus on segmentation and bone age estimation in the literature. While interesting topics, a high level of performance has already been reached and perhaps the attention should be shifted to implementation for these use cases. There is a need for more explainable AI and more methods of explanation over and above case examples and saliency maps.

The quality of research design and reporting varies and guidelines such as those issued by CONSORT and the RSNA [24, 25] need to be followed to improve the quality of research overall.

#### Data

A critical aspect of AI applications in paediatric radiology is the quality and quantity of data used in the studies[26]. As data is the foundation upon which AI algorithms are built and trained, the robustness and generalizability of these algorithms directly depend on the underlying data. In this section, we discuss the quality and quantity of data used in the included studies and the implications for AI model development and performance.

The quality of data in the studies was variable, with some studies employing high-quality, well-curated data from multiple sources, while others relied on single-center, retrospective datasets. Ground truth was determined in various ways, such as radiologic reports, alternative specialist reports (which can reduce reliability [27]), other imaging modalities, or chronological age. The quantity of data in the included studies ranged widely, with a median number of patients of 115 (range 10 - 14,236) and a median number of images of 135 (range 10 - 14,236). The median number of images per patient was only just over. Data were collected from only one hospital in 193 studies (66%), public databases in 39 (13%), and more than one hospital in 32 (11%).

The source code or model was made freely available in only a minority of cases. Greater data sharing, both in terms of code and actual data, can facilitate reproducibility and accelerate the development of robust AI models. The use of data from multiple sources, diverse patient populations, and standardized ground truth determination methods can improve the reliability and generalisability of AI models developed using this data.

#### Bias

The research design and types of data used in studies can introduce various forms of bias, which can impact the validity and generalizability of the results[28–30]. In the context of this survey several biases can arise from the study designs and data types employed [21]:

1. Single-centre effects: When a study is conducted using data from a single centre, the AI model may only learn to recognize patterns and characteristics specific to that center’s population, imaging protocols, and equipment. Consequently, the model’s performance may not generalize well to other centers or broader populations, leading to reduced external validity.
2. Retrospective studies: Where data are collected and analyzed after the events of interest have occurred. This design can introduce various biases, such as selection bias, information bias, and confounding. Furthermore, retrospective studies may not account for potential confounders or variables that could influence the outcome of interest, leading to biased results.
3. Data imbalances: AI models trained on datasets with imbalanced classes (e.g., a disproportionate number of cases from one disease category) can develop a bias towards the majority class. This imbalance can result in a model with poor performance when predicting minority class instances, which can be particularly relevent in paediatric radiology, where certain conditions may be rare.
4. Temporal bias: The performance of AI models may be affected by changes in imaging protocols, equipment, or clinical practice over time. If a study’s data is collected over a long period or does not account for these changes, the AI model may not accurately capture current practices, limiting its applicability to contemporary cases.
5. Ground truth: The determination of ground truth in studies can vary, leading to inconsistencies in the data used to train AI models. For instance, relying solely on radiologic reports may introduce subjectivity and variability, as different radiologists may interpret images differently. In contrast, using alternative sources such as pathology, other imaging modalities, or chronological age may provide more objective ground truth. Inconsistent ground truth can lead to unreliable AI model performance.
6. Evaluation metrics: The choice of evaluation metrics can contribute to bias in the interpretation of an AI model’s performance. Different metrics emphasize different aspects of model performance, which can lead to varying conclusions about the model’s effectiveness. For instance, accuracy is a commonly used metric in classification tasks; however, it may be misleading in imbalanced datasets, as a high accuracy can be achieved by simply predicting the majority class. In such cases, alternative metrics like sensitivity, specificity, F1-score, or area under the receiver operating characteristic curve (AUC-ROC) may provide a more comprehensive understanding of the model’s performance[31].

To mitigate these biases, researchers should consider employing multicentre, prospective study designs with balanced and representative datasets. Additionally, they should ensure consistent ground truth determination methods and account for potential confounders and temporal changes in imaging protocols and clinical practices. By addressing these biases, researchers can develop more reliable and generalizable AI models for paediatric radiology.

## Opportunities

Despite neurology and MSK being the most common subspecialties employing AI (Figure 2), it could be suggested that AI research in paediatric radiology is not purposefully aligned to the most common clinical challenges. Neurodevelopmental disorders and bone age estimation were the most researched use cases. This is in contrast with the most frequently researched and the imaging processing demands in paediatric radiology departments. Imaging procedures in paediatric radiology are mainly requested for cases of trauma, musculoskeletal symptoms or conditions and pulmonary symptoms[32]. In this sense, AI research in paediatric radiology is not currently aligned to the shortage of paediatric radiologists or image interpretation demands in paediatric radiology departments [9]. A similar situation arises by image modality, where the majority of produced images in pediatric radiology being CT and ultrasound but the most researched image modality is MRI[32]. In addition, this is also not contributing to reducing the radiation exposure for children at a population level.

There is a definite need for disruptive solutions for the current supply/demand crisis in radiology. Paediatric radiology presents particular challenges with the diversity and relative scarcity of data but also potential advantages including the motivation of a profound shortage of subspecialists and also the international good-will towards efforts to advance the health of children. Furthermore recent technological improvements have driven down the cost and other barriers to access to high end computing and techniques such as Automated Machine Learning mean that AI is more accessible to clinicians than ever before[33]. Potential for the use of AI in many aspects of radiology workflow have been suggested [22]. Certain paediatric use cases such as paediatric brain tumours or non-accidental injury have the potential to motivate clinicians, computer scientists and engineers alike to translate the potential into outcomes.

## Limitations

This survey paper has limitations, including publication and reporting bias. Studies that do not have a clear methodology may have been misclassified. Furthermore, the heterogeneity of the included studies does not allow for meaningful meta-analysis of results. The high number of included articles only allows for a high-level overview of some themes. Numerous commercial devices utilising AI at varying levels have been introduced. Some of these devices have supporting published literature demonstrating their effectiveness; however, few explicitly disclose details of their underlying algorithms. It is possible that the choice of search terms may have led to the omission of some relevant papers. While we acknowledge that pre-print repositories such as ArXiv are useful for the AI research community as it provides rapid dissemination of novel ideas, they were not included because they are preprints and have not undergone the peer-review process, which ensures the quality and reliability of the research.

## Conclusions

In conclusion, this survey paper provides a comprehensive overview of the advancements in AI literature within paediatric radiology, highlighting the trends in research and the diverse range of subspecialties and modalities involved. Although there has been significant progress, challenges persist in terms of data quality, data sharing, research design and explicability. The present review serves as a broad reference point for researchers. First, it is an index of useful methods to improve patient care. Second, it provides an entry point to enable new collaboration among multidisciplinary teams to tackle paediatric radiology challenges. Finally, it redirects the attention of researchers to areas of inquiry that are currently being overlooked in terms of AI tasks, clinical cases and type of diagnostic images. Tackling this research gaps will aid AI to effectively add value to paediatric radiology.

## Data Availability

All data produced in the present study are available upon reasonable request to the authors

## Online Supplementary Material

Online Supplementary Material 1 - Search terms

Attached

Online Supplementary Material 2- All included papers

Attached

## REFERENCES

1. Langlotz CP (2019) Will Artificial Intelligence Replace Radiologists? Radiology Artif Intell 1:e190058. 10.1148/ryai.2019190058

2. Gillies RJ, Kinahan PE, Hricak H (2016) Radiomics: Images Are More than Pictures, They Are Data. Radiology 278:563–577. 10.1148/radiol.2015151169

3. Thrall JH, Li X, Li Q, et al (2018) Artificial Intelligence and Machine Learning in Radiology: Opportunities, Challenges, Pitfalls, and Criteria for Success. J Am Coll Radiol 15:504–508. 10.1016/j.jacr.2017.12.026

4. Rauschecker AM, Rudie JD, Xie L, et al (2020) Artificial Intelligence System Approaching Neuroradiologist-level Differential Diagnosis Accuracy at Brain MRI. Radiology 295:190283. 10.1148/radiol.2020190283

5. Hosny A, Parmar C, Quackenbush J, et al (2018) Artificial intelligence in radiology. Nat Rev Cancer 18:500–510. 10.1038/s41568-018-0016-5

6. Bluemke DA, Moy L, Bredella MA, et al (2019) Assessing Radiology Research on Artificial Intelligence: A Brief Guide for Authors, Reviewers, and Readers—From the Radiology Editorial Board. Radiology 294:192515. 10.1148/radiol.2019192515

7. Kelly BS, Judge C, Bollard SM, et al (2022) Radiology artificial intelligence: a systematic review and evaluation of methods (RAISE). Eur Radiol 1–10. 10.1007/s00330-022-08784-6

8. Moore MM, Slonimsky E, Long AD, et al (2019) Machine learning concepts, concerns and opportunities for a pediatric radiologist. Pediatr Radiol 49:509–516. 10.1007/s00247-018-4277-7

9. Rimmer A (2017) Radiologist shortage leaves patient care at risk, warns royal college. Bmj 359:j4683. 10.1136/bmj.j4683

10. Roebuck DJ (1999) Risk and benefit in paediatric radiology. Pediatr Radiol 29:637–640. 10.1007/s002470050666

11. Arlachov Y, Ganatra RH (2012) Sedation/anaesthesia in paediatric radiology. Br J Radiology 85:e1018–e1031. 10.1259/bjr/28871143

12. Halabi SS, Prevedello LM, Kalpathy-Cramer J, et al (2019) The RSNA Pediatric Bone Age Machine Learning Challenge. Radiology 290:498–503. 10.1148/radiol.2018180736

13. Keane PA, Topol EJ (2018) With an eye to AI and autonomous diagnosis. Npj Digital Medicine 1:40. 10.1038/s41746-018-0048-y

14. Otjen JP, Stanescu AL, Alessio AM, Parisi MT (2020) Ovarian torsion: developing a machine-learned algorithm for diagnosis. Pediatr Radiol 50:706--714. 10.1007/s00247-019-04601-3

15. Litjens G, Kooi T, Bejnordi BE, et al (2017) A survey on deep learning in medical image analysis. Med Image Anal 42:60–88. 10.1016/j.media.2017.07.005

16. Moher D, Altman DG, Liberati A, Tetzlaff J (2011) PRISMA Statement. Epidemiology 22:128. 10.1097/ede.0b013e3181fe7825

17. Boutron I, Page MJ, Higgins JP, et al (2019) Cochrane Handbook for Systematic Reviews of Interventions. 177–204. 10.1002/9781119536604.ch7

18. Kelly B, Judge C, Bollard SM, et al (2020) Radiology artificial intelligence, a systematic evaluation of methods (RAISE): a systematic review protocol. Insights Imaging 11:133. 10.1186/s13244-020-00929-9

19. Choy G, Khalilzadeh O, Michalski M, et al (2018) Current Applications and Future Impact of Machine Learning in Radiology. Radiology 288:318–328. 10.1148/radiol.2018171820

20. Westgate MJ (2019) revtools: An R package to support article screening for evidence synthesis. Res Synth Methods 10:606–614. 10.1002/jrsm.1374

21. Bs K, C J, S H, et al (2023) How to apply evidence-based practice to the use of artificial intelligence in radiology (EBRAI) using the data algorithm training output (DATO) method. British Journal of Radiology. 10.1259/bjr.20220215

22. Davendralingam N, Sebire NJ, Arthurs OJ, Shelmerdine SC (2021) Artificial intelligence in paediatric radiology: Future opportunities. Br J Radiology 94:20200975. 10.1259/bjr.20200975

23. Otjen JP, Moore MM, Romberg EK, et al (2022) The current and future roles of artificial intelligence in pediatric radiology. Pediatr Radiol 52:2065–2073. 10.1007/s00247-021-05086-9

24. Mongan J, Moy L, Jr CEK (2020) Checklist for Artificial Intelligence in Medical Imaging (CLAIM): A Guide for Authors and Reviewers. Radiology Artif Intell 2:e200029. 10.1148/ryai.2020200029

25. Liu X, Rivera SC, Moher D, et al (2020) Reporting guidelines for clinical trial reports for interventions involving artificial intelligence: the CONSORT-AI extension. Nat Med 26:1364– 1374. 10.1038/s41591-020-1034-x

26. Willemink MJ, Koszek WA, Hardell C, et al (2020) Preparing Medical Imaging Data for Machine Learning. Radiology 295:4–15. 10.1148/radiol.2020192224

27. Luyckx E, Bosmans JML, Broeckx BJG, et al (2019) Radiologists as Co-Authors in Case Reports Containing Radiological Images: Does Their Presence Influence Quality? J Am Coll Radiol 16:526–527. 10.1016/j.jacr.2018.07.035

28. Rouzrokh P, Khosravi B, Faghani S, et al (2022) Mitigating Bias in Radiology Machine Learning: 1. Data Handling. Radiology Artif Intell 4:e210290. 10.1148/ryai.210290

29. Zhang K, Khosravi B, Vahdati S, et al (2022) Mitigating Bias in Radiology Machine Learning: 2. Model Development. Radiology Artif Intell 4:e220010. 10.1148/ryai.220010

30. Faghani S, Khosravi B, Zhang K, et al (2022) Mitigating Bias in Radiology Machine Learning: 3. Performance Metrics. Radiology Artif Intell 4:e220061. 10.1148/ryai.220061

31. Maier-Hein L, Reinke A, Christodoulou E, et al (2022) Metrics reloaded: Pitfalls and recommendations for image analysis validation. Arxiv

32. Tompane T, Bush R, Dansky T, Huang JS (2013) Diagnostic Imaging Studies Performed in Children Over a Nine-Year Period. Pediatrics 131:e45–e52. 10.1542/peds.2012-1228

33. Korot E, Guan Z, Ferraz D, et al (2021) Code-free deep learning for multi-modality medical image classification. Nat Mach Intell 3:288–298. 10.1038/s42256-021-00305-2

