## Supplemental Table 1 for "A survey of Paediatric Radiology Artificial Intelligence"

| **Artificial intelligence** | **Radiology** | **Pediatrics** |
| --- | --- | --- |
| (Artificial intelligence[Title/Abstract])OR  (Machine learning[Title/Abstract])OR  (Support vector machine[Title/Abstract])OR  (SVM[Title/Abstract])OR  (CNN[Title/Abstract])OR  (RNN[Title/Abstract])OR  (LSTM[Title/Abstract])OR  (ResNet[Title/Abstract])OR  (DenseNet[Title/Abstract])OR  (Unet[Title/Abstract])OR  (U-net[Title/Abstract])OR  (DNN[Title/Abstract])OR  (Neural network*[Title/Abstract])OR  (Convolutional network*[Title/Abstract])OR  (Deep learn*[Title/Abstract])OR  (Semantic segmentation[Title/Abstract])OR  (Ensemble[Title/Abstract])OR  (Classification tree[Title/Abstract])OR  (regression tree[Title/Abstract])OR  (probability tree[Title/Abstract])OR  (nearest neighbo*[Title/Abstract])OR  (fuzzy logi*[Title/Abstract])OR  (random forest[Title/Abstract])OR  (kernel[Title/Abstract])OR  (k-means[Title/Abstract])OR  (naive bayes[Title/Abstract]) | (X-ray*[Title/Abstract])OR  (X-ray*[Title/Abstract])OR  (Radiography[Title/Abstract])OR  (Radiograph*[Title/Abstract])OR  (Computed tomography[Title/Abstract])OR  (CT[Title/Abstract])OR  (CAT[Title/Abstract])OR  (CTA[Title/Abstract])OR  (Computerized axial tomography[Title/Abstract])OR  (Magnetic resonance imag*[Title/Abstract])OR  (MRI[Title/Abstract])OR  (MR[Title/Abstract])OR  (Magnetic resonance angio*[Title/Abstract])OR  (MRA[Title/Abstract])OR  (Scintigraphy[Title/Abstract])OR  (DMSA[Title/Abstract])OR  (Ultrasound*[Title/Abstract])OR  (Sonograph*[Title/Abstract])OR  (PET[Title/Abstract])OR  (Positron Emission Tomography[Title/Abstract])OR  (SPECT[Title/Abstract])OR  (Single-photon emission[Title/Abstract])OR  (Single photon emission[Title/Abstract])OR  (mammogra*[Title/Abstract]) | Infan* OR newborn* OR new-born* OR perinat* OR neonat* OR baby OR baby*  OR babies OR toddler* OR minors OR minors* OR boy OR boys OR boyfriend  OR boyhood OR girl* OR kid OR kids OR child OR child* OR children* OR  schoolchild* OR schoolchild OR school child[tiab] OR school child*[tiab] OR  adolescen* OR juvenil* OR youth* OR teen* OR under*age* OR pubescen*  OR pediatrics[mh] OR pediatric* OR paediatric* OR peadiatric* OR school  [tiab] OR school*[tiab] OR prematur* OR preterm* |
